# Epidemiological characteristics of COVID-19 cases in non-Italian nationals notified to the Italian surveillance system

**DOI:** 10.1101/2020.09.22.20199398

**Authors:** Massimo Fabiani, Alberto Mateo-Urdiales, Xanthi Andrianou, Antonino Bella, Martina Del Manso, Stefania Bellino, Maria C. Rota, Stefano Boros, Maria F. Vescio, Fortunato P. D’Ancona, Andrea Siddu, Ornella Punzo, Antonietta Filia, Silvio Brusaferro, Giovanni Rezza, Maria G. Dente, Silvia Declich, Patrizio Pezzotti, Flavia Riccardo, for the COVID-19 working group

## Abstract

**Background:** International literature suggests that disadvantaged groups are at higher risk of morbidity and mortality from SARS-CoV-2 infection due to poorer living/working conditions and barriers to healthcare access. Yet, to date, there is no evidence of this disproportionate impact on non-national individuals, including economic migrants, short-term travellers, and refugees.

**Methods:** We analysed data from the Italian surveillance system of all COVID-19 laboratory-confirmed cases tested positive from the beginning of the outbreak (20^th^ of February) to the 19^th^ of July 2020. We used multilevel negative-binomial regression models to compare the case-fatality rate and the rate of admission to hospital and intensive care unit (ICU) between Italian and non-Italian nationals. The analysis was adjusted for differences in demographic characteristics, pre-existing comorbidities, and period of diagnosis.

**Results:** We analysed 213,180 COVID-19 cases, including 15,974 (7.5%) non-Italian nationals. We found that, compared to Italian cases, non-Italian cases were diagnosed at a later date and were more likely to be hospitalised [(adjusted relative risk (ARR)=1.39, 95% confidence interval (CI): 1.33-1.44)] and admitted to ICU (ARR=1.19, 95% CI: 1.07-1.32), with differences being more pronounced in those coming from countries with lower HDI. We also observed an increased risk of death in non-Italian cases from low-HDI countries (ARR=1.32, 95% CI: 1.01-1.75).

**Conclusions:** A delayed diagnosis in non-Italian cases could explain their worse outcomes compared to Italian cases. Ensuring early access to diagnosis and treatment to non-Italians could facilitate the control of SARS-CoV-2 transmission and improve health outcomes in all people living in Italy, regardless of nationality.

## Introduction

In December 2019, an outbreak of a novel coronavirus (SARS-CoV-2) emerged in China and spread rapidly worldwide, being declared by the World Health Organisation (WHO) a public health emergency of international concern on the 30^th^ of January 2020 and a global pandemic on the 11^th^ of March 2020.^1^ Since then, as of the 23^rd^ of August the pandemic has caused approximately 800,000 deaths globally.^2^

In order to effectively control the pandemic and assess its impact, it is essential to consider how it affects different population groups.

International reports suggest that minority ethnic groups are disproportionally affected by SARS-CoV-2 infection due to cultural, behavioural, and societal characteristics, including disadvantaged socioeconomic conditions, diverse health-seeking behaviour, and intergenerational cohabitation.^3-7^ Some of these aspects may also apply to migrant populations as recent articles have called to consider.^8^ Yet, to date, there is no evidence of this disproportionate impact on this population group. There is evidence, however, that international migrants are more likely to be socioeconomically deprived, to suffer from job insecurity and to be marginalised, all of which are risk factors associated with poor health and poor access to healthcare.^9^ International migrants are also more likely to be employed in low-skilled jobs that have been associated with an increased risk of SARS-CoV-2 infection, such as care work, hospitality, or construction.^10,11^ There is evidence, as well, that migrants face administrative, cultural and language barriers when accessing healthcare.^12,13^

Italy has been one of the most severely affected countries during the first wave of the pandemic. By mid-August 2020, over 250,000 persons had been diagnosed with the infection, and over 35,000 had died from it.^14^ In January 2020, almost 5.3 million foreign citizens were estimated to be living in Italy (8.8% of the total resident population).^15^

On one side, there is concern that non-Italian nationals may be affected differently by the SARS-CoV-2 pandemic compared with the local population, in particular if the barriers described lead to underdiagnosis or to a delayed diagnosis of SARS-CoV-2.

On the other side, however, considering the strong association between COVID-19 related deaths and older age and co-morbidities,^16,17^ it is also possible that foreign persons may be at lower risk of being severely affected by SARS-CoV-2 infection given that, in spite of the higher risk of unhealthy lifestyles associated with disadvantaged socioeconomic conditions, they are generally younger and healthier compared with the local population.^18^ It is also controversial whether Bacillus Calmette-Guérin (BCG) vaccine could have a protective effect against SARS-CoV-2 infection,^19,20^ with possible benefit among foreigners from countries where BCG vaccination is routinely implemented.^21^

In this article, we describe the epidemiology of the SARS-CoV-2 infection among non-Italian cases tested positive in Italy, comparing their distribution over time, case fatality rate (CFR), hospitalisation rate (HR), and rate of admission to intensive care unit (ICU) with those observed in the Italian cases.

## Methods

This study was conducted using routinely collected data and did not rely on a pre-existing analysis plan. We described the methods and presented findings according to the reporting guidelines for observational studies that are based on routinely collected health data (The RECORD statement – checklist of items extended from the STROBE statement) (Supplementary table S1).^22^

### Data sources

We used information retrieved from the Italian national case-base COVID-19 surveillance system established on the 27^th^ of February 2020 and coordinated by the Italian National Institute of Health.^23^ As previously described,^16^ the system collects data on all cases of SARS-CoV-2 infection in Italy laboratory-confirmed by RT-PCR, following the international case definition.^24^ The system collects information on the demographic and clinical characteristics of COVID-19 cases, including reported nationality and outcome of the infection (e.g., hospitalisation, admission to ICU and death). Data is collected and entered daily by the 19 regions and the two autonomous provinces using a secure online platform. Data is checked for out-of-range values, inconsistencies, and duplicated records at the coordinating centre. A list of possible errors in data-entry is routinely sent to regions for verification and possible corrections. For the purpose of this study, we used the complete dataset of notifications updated on the 25^th^ of August 2020 and selected all COVID-19 cases tested positive in Italy from the 20^th^ of February to the 19^th^ of July 2020, thus accounting for at least 30 days of follow-up since the date of testing and a week of possible delay in information updating.

The scientific dissemination of COVID-19 surveillance data was authorised by the Italian Presidency of the Council of Ministers on the 27^th^ of February 2020 (Ordinance n. 640).

### Outcomes, exposure, and confounders

Among all notified COVID-19 cases, we defined as non-Italian nationals all people with a reported non-Italian nationality, regardless of citizenship or country of birth. We classified the nationality as Italian vs. non-Italian in general and according to the 2018 human development index (HDI) of the country of origin.^25^ To this purpose we defined three categories based on the tertiles of the world’s countries HDI distribution (low-HDI country: HDI ≤ 0.651; medium-HDI country: 0.651 < HDI < 0.799; high-HDI country: ≥ 0.799).

We analysed the associations between nationality (exposure) and different outcomes. First, we assessed and compared the CFR, both overall and among hospitalised cases, considering COVID-19 associated deaths any notified person who was confirmed to be infected with SARS-CoV-2 and has died within 30 days since the date of testing positive. Second, within the same time-interval, we assessed and compared the HR and, among cases who were hospitalised, the rate of admission to ICU (ICUR).

We considered as potential confounders of the relationship between nationality and outcomes the following variables: sex, age (categorised as < 30 years of age, 5-years age groups from 30-34 years to 60-74 years of age, and ≥ 75 years of age), Italian geographical macro-area of diagnosis (i.e., North-West, North-East, Centre, and South and Islands), level of urbanisation of the place of residence (i.e., urban, semi-urban and rural), pre-existing comorbidities (i.e.. presence of at least one of oncologic, cardiovascular, respiratory, and diabetes/metabolic diseases), and calendar period of testing (i.e., approximately 30-day time intervals from 20/February-19/March to 20/June-19/July).

### Statistical analysis

We excluded from the analysis all cases presenting data inconsistencies (i.e., death occurred before hospitalisation), cases who were imported from abroad, and those with missing information about nationality. Moreover, we excluded from the multivariable analysis all cases with missing information about age, level of urbanisation of the residence area, and pre-existing comorbidities (see figure 1).

**Figure 1.**
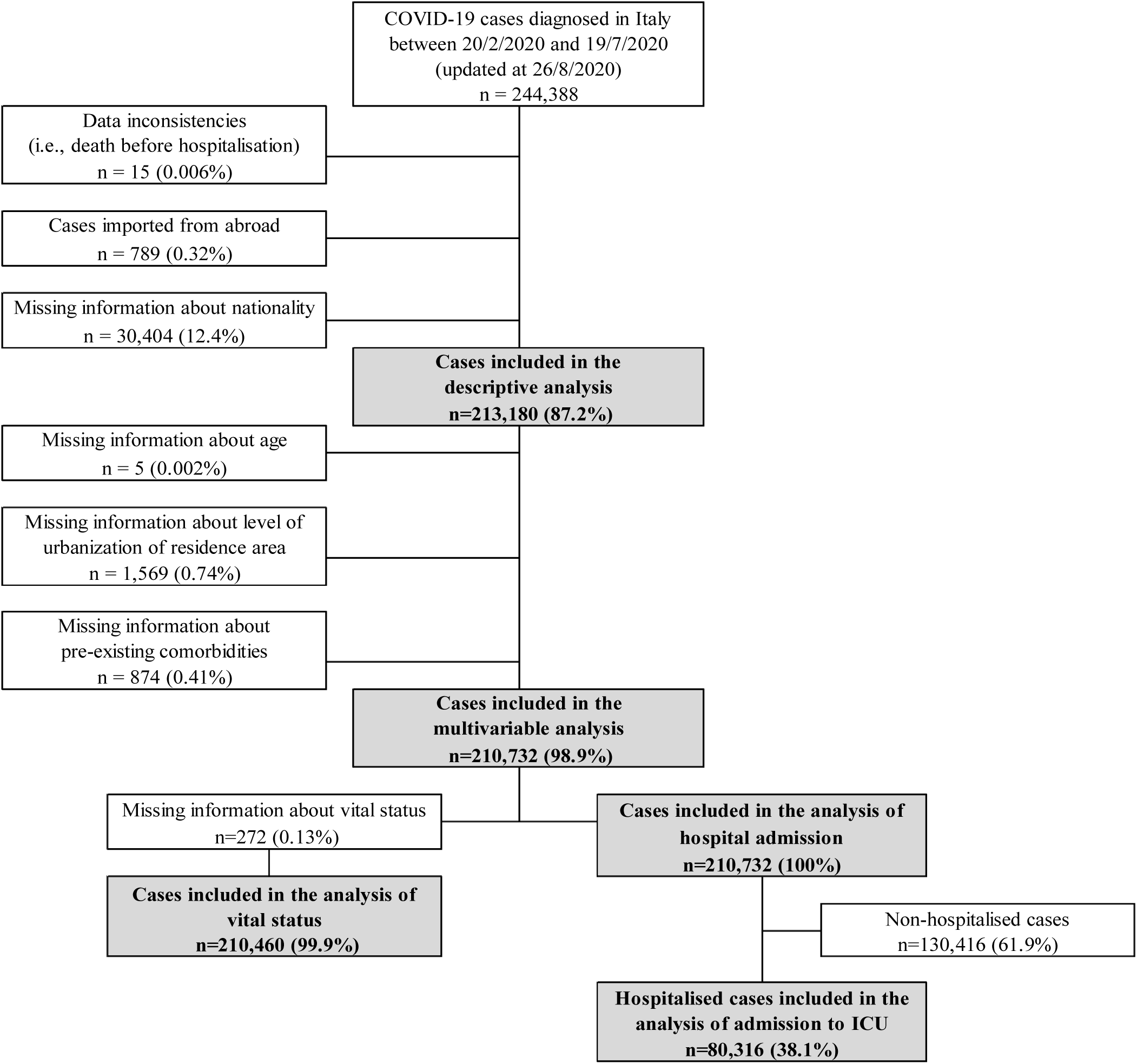
Selection of COVID-19 cases included in the study. ICU, intensive care unit.

**Figure 3.**
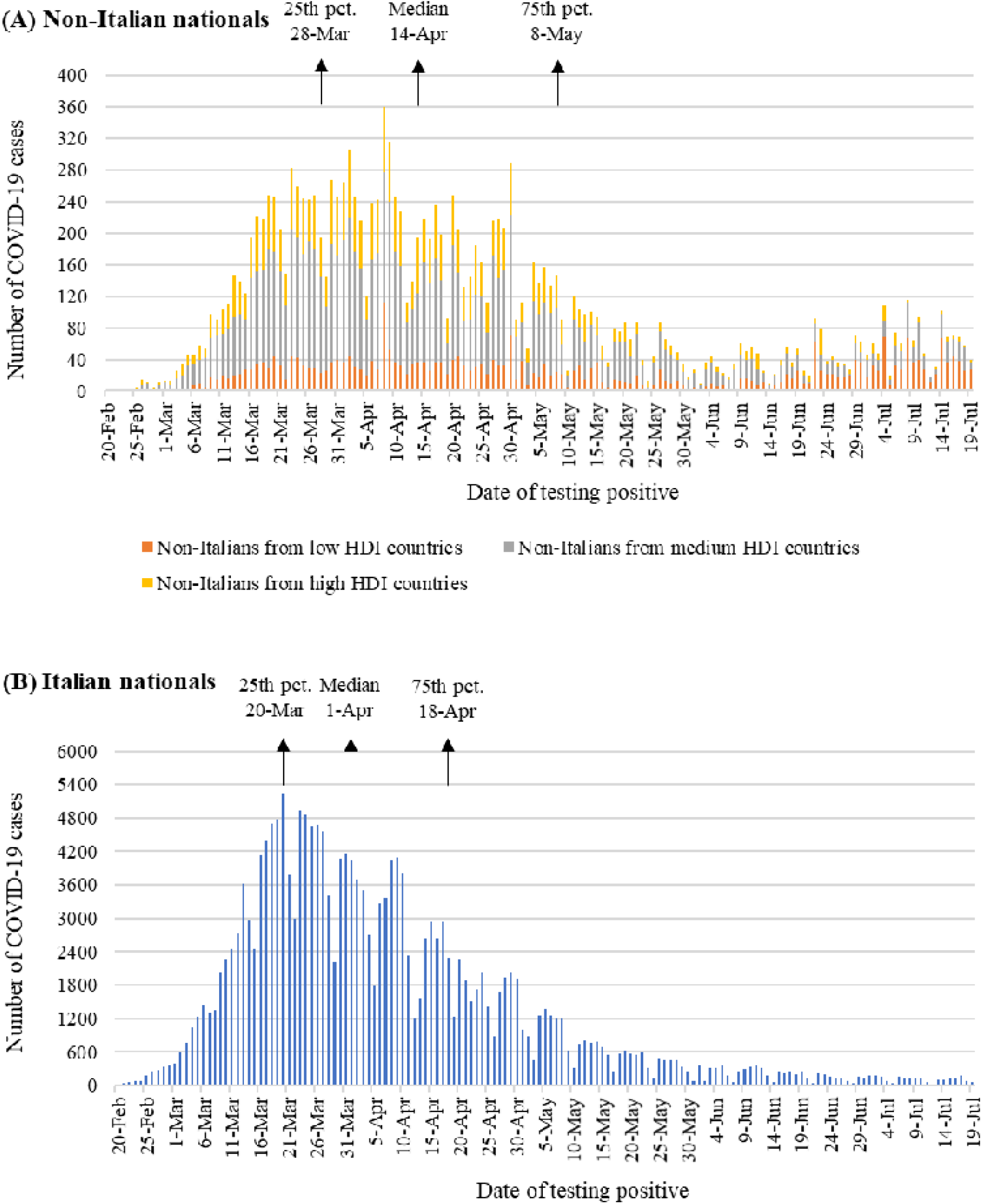
COVID-19 epidemiological curves in (A) non-Italian nationals (B) and Italian nationals, 20 Feb. – 19 July 2020. HDI, human development index. Pct., percentile.

We described the main demographic and clinical characteristics and the distribution of cases over time by nationality using counts with percentages and median with interquartile range (IQR) for categorical and continuous variables, respectively. Differences were evaluated using the chi-square test (categorical variables) or the Wilcoxon rank-sum test or Kruskall-Wallis test (continuous variables).

We used multilevel negative-binomial regression models to estimate the adjusted CFR ratios (CFRR), HR ratios (HRR), and ICUR ratios (ICURR) for non-Italian cases compared to Italian cases. Rate ratios were adjusted for sex, age group, geographical macro-area of diagnosis, level of urbanisation of the place of residence, pre-existing comorbidities, and calendar period of diagnosis. A random effect accounting for regional differences in health contexts and strategies was also included into the models. All estimates were presented together with their 95% confidence interval (CI).

We tested possible interactions between nationality and each covariate included into the multivariable models through the likelihood-ratio test.

Finally, to evaluate a possible selection bias due to missing information about nationality, we conducted a sensitivity analysis assuming that cases with no information about nationality but with a fiscal code indicative that they were born abroad or in Italy were non-Italian nationals or Italian nationals, respectively.

All tests were two-sided and statistical significance was set at p < 0.05. The analyses were performed using Stata/SE version 16.0 (StataCorp LLC, Texas, USA).

## Results

After the exclusion of cases presenting data inconsistencies [15 (0.006%)], cases imported from abroad [789 (0.32%)], and those with missing information about nationality [30,404 (12.4%)], a total of 213,180 (87.2%) out of the 244,388 initial cases were included in the analysis (see figure 1). Of these, 15,974 were non-Italian nationals (7.5%) and 197,206 were Italian nationals (92.5%). Among the non-Italian nationals, almost all had an Italian fiscal code indicating that they were regular migrants [15,546 (97.3%)].

### Socio-demographic characteristics and time of diagnosis

According to the distribution by world’s macro-area presented in Supplementary figure S1, cases of COVID-19 among non-Italian nationals occurred in people from various countries. Non-Italian cases from low-HDI countries were almost all from south-central Africa [1,834 (53.6%)] and Asia [1,553 (45.4%)]. Non-Italian cases from medium-HDI countries were mainly from south-central America [3,551 (42.0%)], European countries outside the European Union (EU) [2,606 (30.8%)], and North Africa [1,517 (18.0%)], while non-Italians from high-HDI countries were in large part from other EU countries [3,447 (84.0%)].

Among all COVID-19 cases, compared with Italian nationals [91,376 males (46.3%)], non-Italian nationals were less frequently males [6,871 (43.0%)] (p < 0.001), especially those from high-HDI countries [1,150 (28.0%)], while the difference was reversed in non-Italian nationals from low-HDI countries [2,372 (69.3%)] (Table 1). All cases among non-Italian nationals (median age: 44 years; IQR: 35-53) were younger than Italian cases (median age: 63 years; IQR: 48-80) (p < 0.001), especially those from low-HDI countries (median age: 37 years; IQR: 28-46). Non-Italian nationals were more frequently diagnosed in northern Italy and residing in urban areas [9,662 (60.5%) and 8,172 (52.0%), respectively] compared to Italian nationals [106,276 (53.9%) and 64,802 (33.1%), respectively] (p < 0.001). This difference was more evident among non-Italian nationals from medium-HDI countries [5,668 (67.1%) and 4,764 (57.1%), respectively].

**Table 1.**
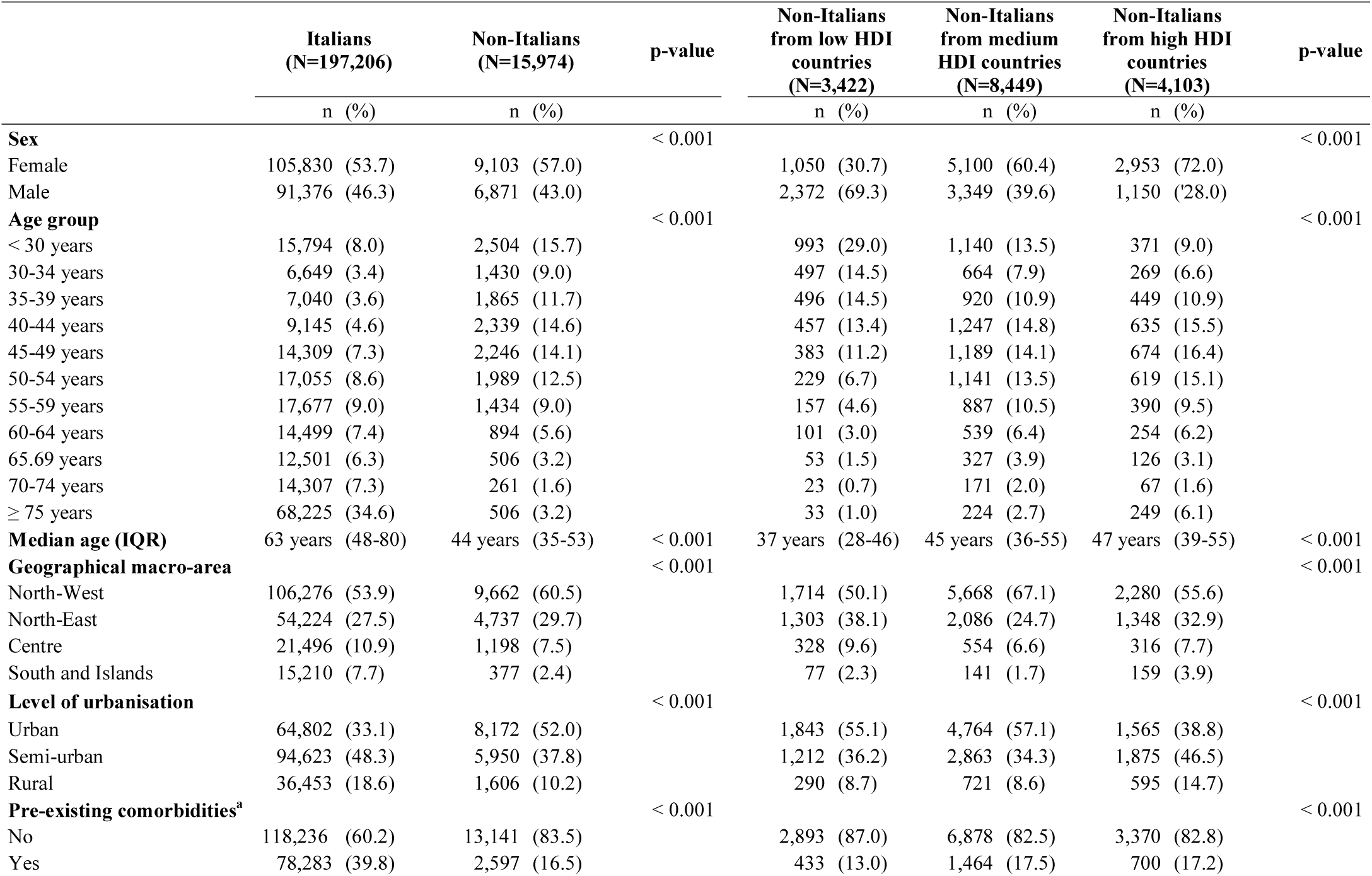
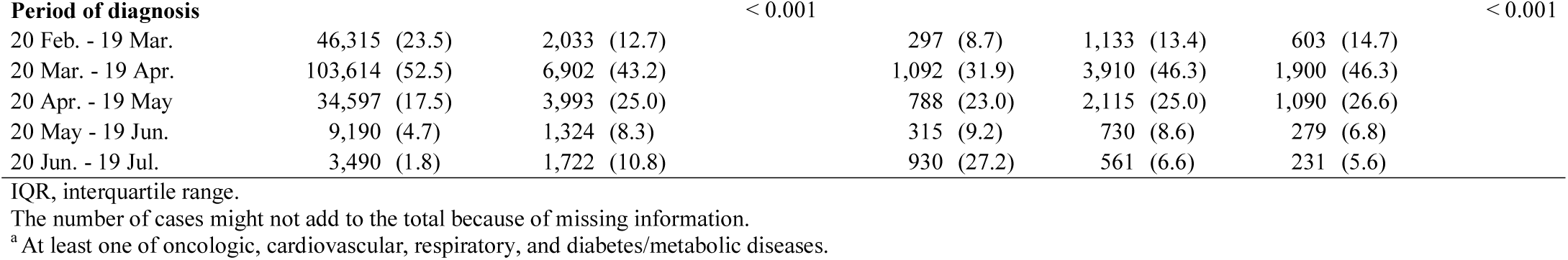
Demographic and clinical characteristics of the COVID-19 cases included in the study.

Pre-existing comorbidities were less frequently reported in non-Italian cases [2597 (16.5%)] compared to Italian cases [78,283 (39.8%)] (p < 0.001), especially in those from low-HDI countries [433 (13.0%)].

In general, non-Italian cases were diagnosed on a later date compared with Italian cases (see figure 2). The median date of testing positive was 14/April (IQR: 28/March – 8/May) for non-Italian nationals compared to 1/April (IQR: 20/March – 18/April) for Italian nationals (p < 0.001). This difference was particularly evident among non-Italian nationals from low-HDI countries (median date at diagnosis: 29/April; IQR: 6/April – 22/June).

### Case fatality rate, hospitalisation rate, and rate of admission to intensive care unit

Table 2 shows the crude and adjusted CFRRs, HRRs, and ICURRs. Overall, we observed no differences in CFR between non-Italian nationals and Italian nationals. The only exception was for non-Italian nationals from low-HDI countries, who showed an increased risk of death compared to Italian nationals (Adj. CFR=1.32, 95% CI: 1.01-1.75), although this difference was not observed among hospitalised cases.

**Table 2.** Case fatality rate, hospitalization rate, rate of admission to intensive care units, and in-hospital lethality in non-Italian nationals compared to Italian nationals.

| Case fatality rate (CFR) | Cases | Deaths | CFR (%) | Crude CFRR | (95% CI) | Adj. CFRR<sup>a</sup> | (95% CI) |
| --- | --- | --- | --- | --- | --- | --- | --- |
| **Nationality (1)** |  |  |  |  |  |  |  |
| Italian | 194,956 | 27,836 | 14.3 | ref. |  | ref. |  |
| Non-Italian | 15,504 | 390 | 2.5 | 0.18 | (0.16-0.19) | 1.01 | (0.91-1.14) |
| **Nationality (2)** |  |  |  |  |  |  |  |
| Italian | 194,956 | 27,836 | 14.3 | ref. |  | ref. |  |
| Low HDI countries | 3,255 | 53 | 1.6 | 0.11 | (0.09-0.15) | 1.32 | (1.01-1.75) |
| Medium HDI countries | 8,246 | 237 | 2.9 | 0.20 | (0.18-0.23) | 1.03 | (0.89-1.18) |
| High HDI countries | 4,003 | 100 | 2.5 | 0.17 | (0.14-0.21) | 0.85 | (0.69-1.04) |
| Hospitalisation rate (HR) | Cases | Hospitalised | HR (%) | Crude HRR | (95% CI) | Adj. HRR<sup>1</sup> | (95% CI) |
| **Nationality (1)** |  |  |  |  |  |  |  |
| Italian | 195,225 | 75,432 | 38.6 | ref. |  | ref. |  |
| Non-Italian | 15,507 | 4,884 | 31.5 | 0.82 | (0.79-0.84) | 1.39 | (1.33-1.44) |
| **Nationality (2)** |  |  |  |  |  |  |  |
| Italian | 195,225 | 75,432 | 38.6 | ref. |  | ref. |  |
| Low HDI countries | 3,255 | 925 | 28.4 | 0.74 | (0.69-0.78) | 1.59 | (1.48-1.71) |
| Medium HDI countries | 8,249 | 2,970 | 36.0 | 0.93 | (0.90-0.97) | 1.46 | (1.39-1.53) |
| High HDI countries | 4,003 | 989 | 24.7 | 0.64 | (0.60-0.68) | 1.15 | (1.07-1.23) |
| Rate of admission to ICU (ICUR) in hospital. cases | Hospitalised cases | Admitted to ICU | ICUR (%) | Crude ICURR | (95% CI) | Adj. ICURR<sup>1</sup> | (95% CI) |
| **Nationality (1)** |  |  |  |  |  |  |  |
| Italian | 75,432 | 9,066 | 12.0 | ref. |  | ref. |  |
| Non-Italian | 4,884 | 549 | 11.2 | 0.94 | (0.86-1.02) | 1.19 | (1.07-1.32) |
| **Nationality (2)** |  |  |  |  |  |  |  |
| Italian | 75,432 | 9,066 | 12.0 | ref. |  | ref. |  |
| Low HDI countries | 925 | 92 | 9.9 | 0.83 | (0.67-1.02) | 1.16 | (0.93-1.44) |
| Medium HDI countries | 2,970 | 354 | 11.9 | 0.99 | (0.89-1.10) | 1.25 | (1.11-1.42) |
| High HDI countries | 989 | 103 | 10.4 | 0.87 | (0.71-1.05) | 1.07 | (0.88-1.32) |
| Case fatality rate (CFR) in hospital. cases | Hospitalised cases | Deaths | CFR (%) | Crude CFRR | (95% CI) | Adj. CFRR<sup>1</sup> | (95% CI) |
| **Nationality (1)** |  |  |  |  |  |  |  |
| Italian | 75,252 | 20,823 | 27.7 | ref. |  | ref. |  |
| Non-Italian | 4,883 | 327 | 6.7 | 0.24 | (0.22-0.27) | 0.93 | (0.83-1.04) |
| **Nationality (2)** |  |  |  |  |  |  |  |
| Italian | 75,252 | 20,823 | 27.7 | ref. |  | ref. |  |
| Low HDI countries | 925 | 37 | 4.0 | 0.14 | (0.10-0.20) | 0.93 | (0.67-1.29) |
| Medium HDI countries | 2,969 | 206 | 6.9 | 0.25 | (0.22-0.29) | 0.93 | (0.80-1.07) |
| High HDI countries | 989 | 84 | 8.5 | 0.31 | (0.25-0.38) | 0.94 | (0.76-1.17) |

ARR, AR ratio; CFRR, CFR ratio; HRR, HR ratio, ICU, intensive care unit, ICURR, ICUR ratio; CI, confidence interval; ref., reference category.
<sup>a</sup> Adjusted for sex, age group, geographical macro-area of diagnosis, level of urbanisation of the place of residence, pre-existing comorbidities, and calendar period of diagnosis, and accounting for the random effect due regional contextual differences.

Moreover, we found that HR was significantly higher in cases among non-Italian nationals (Adj. HRR=1.39, 95% CI: 1.33-1.34), especially in those from low-HDI countries (Adj. HRR=1.59, 95% CI: 1.48-1.71).

Finally, we found that ICUR among hospitalised cases was also higher in non-Italian nationals compared to Italian nationals (Adj. ICURR=1.19, 95% CI: 1.07-1.32), particularly in those from medium-HDI countries (Adj. ICURR=1.25, 95% CI: 1.11-1.42) and low-HDI countries (Adj. ICURR=1.16, 95% CI: 0.93-1.44).

With reference to the risk of hospitalisation, we found significant interactions between nationality and sex (p=0.001), age group (p < 0.001), geographical macro-area of diagnosis (p < 0.001), presence of pre-existing comorbidities (p < 0.001), and calendar period of diagnosis (p < 0.001). However, stratum-specific estimates of HRR for non-Italian cases compared to Italian cases were generally in the same direction as in the main model (detailed results from models with interaction terms are presented in Supplementary table S4).

## Discussion

The results show that, at this stage of the COVID-19 epidemic in Italy, non-Italian cases were more likely to be hospitalised and to be admitted to ICU compared with Italian cases, also showing a higher probability to die from the infection in those from low-HDI countries. We observed an inverse gradient by which the risk of hospitalisation, admission to ICU, and death increased as the human development index of the country of origin decreased.

The epidemic curves of non-Italian nationals appear shifted to the right, indicating a median date of diagnosis approximately two weeks later compared with that of Italian nationals (four weeks later among non-Italian nationals from low-HDI countries). This suggests that non-Italian cases were diagnosed in a less timely fashion than Italian cases, possibly when the disease was more advanced and symptoms more severe. A seroprevalence study of SARS-CoV-2 conducted in north-eastern Italy corroborates this hypothesis, showing that immigrant women were more likely to be tested only when their symptoms were severe.^26^ Moreover, a delay of diagnosis in the foreign population has been previously described for other infections in Italy, such as tuberculosis.^27^ The delay in diagnosis, likely associated with worse clinical conditions, could explain the increased rate of hospitalisation, admission to ICU, and death that we observed among non-Italian nationals compared to Italian nationals, especially in those from low-HDI countries.

In Italy, all non-Italian nationals are granted to access emergency services and some out-patient services,^28^ while access to additional services, including being assigned to a general practitioner, the most likely mediator for early diagnosis, occurs only when non-Italian nationals live in Italy with a documented status. However, regardless of status, informal barriers (language, administrative, legal, cultural, and social) hinder the prompt access to healthcare services, likely leading to a delayed diagnosis.^12,13^ In the pandemic context, non-Italian nationals also might have delayed diagnosis fearing restriction of working activities due to isolation/quarantine.^29^ These hypotheses could partly explain the less pronounced decline in incidence observed after the start of the lockdown measures in non-Italian nationals compared to Italian-nationals. In fact, the circulation of the virus in the foreign community could have been higher because of non-Italian cases who were diagnosed late and were not promptly isolated/quarantined at an earlier stage of the disease.

Differences in the epidemic curves could also be explained by a different pace in the penetration of the disease in Italian and non-Italian communities. It is possible that areas with low percentage of non-Italian nationals were affected at an earlier stage, thus shifting the curve of non-Italian cases to the right. However, in that case, under the assumption that the median survival time after diagnosis was the same in the two groups, we would expect a similar shift in the distribution of deaths over time. Although we did observe a delay in the median date of death of non-Italians compared with Italians (4 days), considering that the median survival time of non-Italian dead cases was 2 days higher than that of their Italian counterpart, the magnitude of this delay was 85% smaller compared with the delay in diagnosis (2 days vs 13 days). This suggests that a different pace of penetration of the disease influenced the shift only partially.

Finally, looking at the epidemic curve, we observed a new increase in the number of non-Italians tested positive during the last time-period (20/June – 19/July), possibly explained by an increase in the number of cases imported from abroad. In fact, a large part of the imported cases notified to the surveillance system were tested positive in the last period (79%), most of them in non-Italian nationals (77%). Although we excluded from our analysis all cases imported from abroad, part of them could have not been promptly detected and quarantined at their arrival to Italy and might have favoured the circulation of the virus within the foreign community.

Our results do not support previous findings around the hypothetical protective effect of the BCG vaccination against severe infection and death from SARS-CoV-2,^21^ as we found an increased risk of hospitalisation and admission to ICU in non-Italian cases, including those from countries were BCG is part of the vaccination schedule.

This is, to the best of our knowledge, the first analysis that compares the epidemiology of the SARS-CoV-2 infection in the non-national population using individual data. It is based on information collected from the Italian integrated COVID-19 surveillance system, which covers the entire national territory.

However, this analysis has also some limitations. Unfortunately, we were not able to retrieve reliable estimates of the population size at-risk of infection because the last available population estimates were updated on the 1st of January 2020. Therefore, these estimates do not account for possible population movement occurred in the foreign population, especially during the early phase of the epidemic. This did not consent to accurately estimate and compare the attack rate of SARS-Cov-2 infection between non-Italian nationals and Italian nationals.

Other limitations are mainly related to completeness of surveillance data. We were not able to retrieve information about nationality for 12% of all cases notified during the study period and this could have introduced a bias in our estimates. We conducted a sensitivity analysis assuming that cases with no information about nationality but with a fiscal code indicative that they were born abroad or in Italy were non-Italian nationals or Italian nationals, respectively. Based on this assumption, we were able to classify 3,189 cases as non-Italians and 24,793 cases as Italians out of the 28,942 cases excluded from the analysis. We found that the direction and magnitude of the associations between nationality and all the analysed outcomes were substantially the same as those presented in the results section, thus suggesting that this kind of bias was reduced to a minimum (detailed results of sensitivity analysis are presented in Supplementary table S6).

Finally, we were not able to obtain a reliable estimate of the date of onset of symptoms for cases who were symptomatic at the time of diagnosis because the information was missing for 21% of them. We think, however, that using the difference between the self-reported date of symptom onset and the date of diagnosis to assess possible delay in diagnosis could have been misleading because this information may be strongly affected by a desirability bias. As the social and cultural factors that could influence people’s fear of being stigmatised and punished for reporting social interactions while symptomatic (e.g. for work purposes) may be different in nationals compared with non-nationals, we felt that including this analysis would, therefore, also have been problematic.^30^ In conclusion, our results suggest that, compared to Italian cases, non-Italian cases are more likely to experience a delayed access to health services. Accessing care when the disease is more advanced and clinical conditions possibly more severe can explain why it is more likely for non-Italian nationals to require hospitalisation and admission to ICU. Compared to Italian cases, differences in clinical outcomes appear more pronounced in non-Italian cases from countries with lower HDI and reduced in those from high-HDI countries. Ensuring that all national disease prevention, diagnosis, treatment and control measures are made more accessible to all foreigners, also through enhanced and targeted communication strategies, could facilitate earlier detection of new infections and case management as well as the isolation of cases and the tracing, quarantine, and monitoring of their close contacts. Removing healthcare access barriers and reinforcing communication are, therefore, essential to control SARS-CoV-2 transmission, preserve health services, and improve the health outcomes of all people living in the same country, regardless of nationality.

## Data Availability

Because of data sharing restrictions, the file containing the anonymised dataset cannot be made publicly available.

## Acknowledgements

We thank Dr Aldo Rosano for his helpful support.

## Key points

- We analysed data collected in the framework of the COVID-19 Italian integrated surveillance system, including cases diagnosed in the whole country from the beginning of the local epidemic on the 20^th^ of February to the 19^th^ of July 2020.
- Compared to Italian cases, we found that non-Italian cases were more likely to be hospitalised and admitted to an intensive care unit, also showing an increased probability to die from SARS-CoV-2 infection in those from countries with a low human development index.
- We observed an inverse gradient by which the risk of hospitalisation, admission to intensive care unit, and death increased as the human development index of the country of origin decreased.
- Over the whole study period, non-Italian cases were diagnosed approximately two weeks later compared to Italian cases, thus suggesting that a delayed diagnosis possibly led to worse outcomes.
- If non-nationals are hindered in accessing healthcare services in a timely manner, testing, tracking, and tracing policies can be in turn delayed with a possible negative impact on individual outcome as well as on disease prevention and control at population level.

